# Biomarkers of neurodegeneration in schizophrenia: Systematic Review and Meta-Analysis

**DOI:** 10.1101/2023.10.31.23297823

**Authors:** Jack C. Wilson, Kathy Y. Liu, Katherine Jones, Jansher Mahmood, Utkarsh Arya, Robert Howard

## Abstract

**Question:** Does neurodegenerative disease underlie the increased rate of dementia observed in older people with schizophrenia? Several studies have reported a higher prevalence of dementia in people with schizophrenia compared to the general population. This may reflect higher risk of developing neurodegenerative diseases such as vascular dementia or Alzheimer’s disease (AD). Alternatively, this may reflect nonpathological, age-related cognitive decline in a population with low cognitive reserve. We reviewed the literature on neurodegeneration markers in older people with schizophrenia and dementia or cognitive impairment to establish whether neurodegenerative disease underlies the increased rate of dementia observed.

**Study Selection and Analysis:** We reviewed papers that compared post-mortem findings, hippocampal volume, or CSF markers of AD, in schizophrenia patients with evidence of cognitive impairment (age ≥45 years) with controls. Most studies investigated AD neuropathology. We subsequently performed a meta-analysis of post-mortem studies that compared amyloid-β plaques (APs) or neurofibrillary tangles (NFTs) in cognitively impaired schizophrenia patients to either controls or an AD group.

**Findings:** No studies found significant increase of amyloid-β plaques (APs) or neurofibrillary tangles (NFTs) in cognitively impaired schizophrenia patients compared to controls. All postmortem studies that compared APs or NFTs in schizophrenia patients to an AD group found significantly more APs or NFTs in AD. No studies found significant differences in CSF total tau or phosphorylated tau between schizophrenia patients and controls. Two studies found significantly decreased CSF Aβ42 in schizophrenia compared to patients. Findings for hippocampal volume were mixed.

**Conclusions:** Studies have not found higher rates of AD-related pathology in cognitively impaired schizophrenia individuals compared to controls. Higher rates of dementia identified in population studies may reflect lack of specificity in clinical diagnostic tools used to diagnose dementia.

## Key Messages

### What is already known on this topic

Several large epidemiological studies have identified apparently higher rates of Alzheimer’s disease and dementia in the schizophrenia population. No previous study has reviewed whether established markers of neurodegeneration are present in cognitively impaired people with schizophrenia.

### What this study adds

We found an absence of excess of such markers, casting doubt on whether common neurodegenerative conditions, such as Alzheimer’s disease, truly underlie reported higher rates of dementia in the schizophrenia population.

### How this study might affect research, practice or policy

Higher rates of dementia in schizophrenia may be mainly related to a lack of specificity in dementia diagnostic tests and the possibility that age-related cognitive decline in a group with a low cognitive reserve can cause a diagnostic threshold for dementia to be crossed earlier.

## Introduction

Several studies over recent years have reported increased risk of dementia in people with schizophrenia compared to the general population and those with other psychiatric diagnoses.[1–5] However, the mechanisms behind this are unclear, and it is not known whether specific neurodegenerative disease processes are involved. The hallmark of dementia is *progressive* cognitive decline; i.e. worsening and irreversible symptoms, manifesting over months to years, and beyond what might be expected due to ageing. Cognitive impairment in schizophrenia is well established and observed throughout the course of the illness and, despite heterogeneous trajectories, is believed to remain relatively stable in most cases.[6–8] There are a paucity of longitudinal cognitive data in older people with chronic schizophrenia, which may also be influenced by the effects of institutionalisation.[9] It would be important to determine whether an observed increased risk of dementia can be accounted for by identified neurodegenerative disease processes, otherwise researchers and clinicians risk misclassifying cognitive symptoms of schizophrenia in older people as comorbid dementia.

There are plausible reasons why schizophrenia may truly be associated with higher rates of dementia. Known dementia risk factors, such as smoking, obesity and diabetes, are higher in this population,[10, 11] and average educational attainment and consequent cognitive reserve are lower.[12] There may also be overlapping genetic risk for schizophrenia and neurodegenerative disorders.[13, 14, 15].

Alternatively, apparent higher dementia rates in schizophrenia may be an artefact of assessing older individuals who are experiencing earlier or accelerated age-related cognitive decline,[12] but, due to low cognitive reserve, have crossed a clinical threshold that warrants a dementia diagnosis. Lower scores on cognitive testing are a key component of diagnosing dementia; patients with schizophrenia may perform poorly on these tests due to deficits in attention, working memory, verbal fluency and executive functioning, which are inherent to schizophrenia, as well as the effects of associated drug treatment,[16,17] and institutionalisation,[9] rather than underlying neurodegeneration.[18].

Evidence of neurodegeneration, for example Alzheimer’s disease (AD) pathology, in older schizophrenia patients with cognitive impairment or a diagnosis of dementia would support the proposal that increased neurodegeneration underlies higher risk of developing dementia. Alternatively, if rates of markers of neurodegeneration in this group mirror those in the general population, the increased dementia rates that have been reported in the schizophrenia population must have an alternative explanation. Although an earlier study did not find post-mortem evidence of Alzheimer’s disease in cognitively impaired older individuals with schizophrenia,[19] there has been no systematic assessment of the findings and quality of those studies that have investigated any (including histopathological, fluid and neuroimaging) markers of common neurodegenerative conditions in this population.

We aimed to review the literature on neuropathological and neurodegeneration biomarker studies in older people with schizophrenia, who showed evidence of cognitive impairment or had a clinical diagnosis of dementia, with a view to establishing whether neurodegenerative disease processes underlie the increased rate of dementia observed in this population. A secondary aim was to synthesise the findings of post-mortem studies in a meta-analysis where the required data was available.

## Methods

### Literature search

Online literature databases (Pubmed (from 1964), Web of Science (from 1900) and Embase (from 1974)) were initially searched up to 29th October 2021, and subsequently updated on 31^st^ January 2023, using the search terms schizophreni* OR hebephrenia OR ‘dementia praecox’) AND (dementia OR neurodegenerati* OR Alzheimer* OR ‘cognitive impairment’ OR ‘cognitive dysfunction’ OR ‘cognitive decline’ OR ‘cognition disorders’ OR ‘mild cognitive impairment’) AND (neuropatholog* OR patholog* OR post-mortem OR postmortem OR autopsy OR amyloid OR tau OR ‘neurofilament light chain’ OR hippocamp*) AND (older OR elder* OR ‘old age’ OR ‘geriatric’ OR aging OR ageing OR ‘increasing age’ OR ‘middle age*’ OR ‘middle-age* OR aged OR mid-life).

### Inclusion / exclusion criteria

Studies were included if they were published, peer-reviewed, observational articles in English on human subjects aged >=45 years with a diagnosis of schizophrenia. In addition, subjects had either a comorbid diagnosis of dementia or evidence of cognitive impairment on quantitative neuropsychological tests. Studies were included if they investigated neurodegeneration markers and included a comparison group (i.e., was a case-control study). Neurodegeneration markers were defined as one or more of the following:

– Amyloid-β plaques, abnormal tau/neurofibrillary tangles (NFTs), cerebral amyloid angiopathy, TDP-43, hippocampal sclerosis, Lewy bodies, or vascular pathologies such as infarcts, atherosclerosis, arteriolar sclerosis, identified in post-mortem analyses
– Any other marker not mentioned above, which was directly compared between individuals with schizophrenia versus a neurodegenerative condition, including AD, Parkinson’s disease (PD), Lewy body dementia (DLB), and vascular dementia
– Amyloid or tau, quantified using positron emission tomography (PET), cerebrospinal fluid (CSF), or plasma measures
– Neurofilament light chain analysis in CSF
– Hippocampal volume analysis using MRI

We excluded post-mortem studies that only assessed changes in neurotransmitter systems, or neuropathological changes not associated with neurodegeneration/neurodegenerative disorders (unless they were direct comparisons between schizophrenia and a neurodegenerative group, as above). We also excluded studies that examined other psychotic illnesses, such as bipolar disorder or schizophrenia spectrum disorders (i.e., schizotypal personality disorder, schizoaffective disorder, schizophreniform disorder), and very late-onset schizophrenia-like psychosis, and studies that grouped schizophrenia patients with these disorders if it was not possible to isolate results for the schizophrenia patients. Clinical trials, reviews, case studies, conference abstracts or book chapters were also excluded.

### Data Extraction

Two authors (out of UA, JW, KJ, JM, and KL) independently screened all papers for inclusion based on their titles and abstracts. Two authors (out of UA, JW, KJ and JM) then independently assessed the remaining full text articles for eligibility and performed data extraction on the included papers. Any discrepancies were discussed and re-examined by the two authors and a third author was consulted (KL) if required. We recorded what type of neuropathological data was investigated, differences found between groups, and the conclusions of the authors regarding the prevalence of dementia in schizophrenia, or the underlying aetiology of cognitive impairment in patients with schizophrenia. An assessment of the quality of included studies was also conducted using a modified version of the NIH Quality Assessment Tool for Observational Cohort and cross sectional studies (two questions were omitted due to lack of relevance to our study: *For exposures that can vary in amount or level, did the study examine different levels of the exposure as related to the outcome* and *was the exposure assessed more than once over time?*) Studies were given a score of between 0 and 12, with 0-4 classed as low, 5-8 classed as medium, and 9-12 classed as High.

### Meta Analysis

Following our systematic review of the literature, we performed a meta-analysis on suitable studies. For studies to be suitable for meta-analysis they needed to compare results for either the number of amyloid plaques or neurofibrillary tangles (or both) between cognitively impaired patients with schizophrenia and normal elderly controls or an Alzheimer’s control group (or both). Studies must have provided quantitative results with either an identifiable standard deviation or standard error. Where studies provided AP counts or NFT counts from different areas of the brain, we preferentially selected results from the hippocampus (present in the majority of studies), followed by the neocortex, followed by subcortical structures.

Suitable studies were included in a random effects meta-analysis model to compute a weighted effect size and confidence interval across studies using the ‘dmetar’ package (version 0.0.9000) in R version 4.3.1. Effect size (Hedge’s G) was calculated and results for the four comparisons (APs in schizophrenia patients vs controls, APs in schizophrenia patients vs Alzheimer’s patients, NFTs in schizophrenia patients vs controls, NFTs in schizophrenia patients vs Alzheimer’s patients) were represented as forest plots. Study heterogeneity was measured using the I2 statistic. 95% confidence intervals and the pooled effect size were calculated using the ‘dmetar’ package.

## Results

### Identification and characteristics of included studies

Initial literature searches identified 1696 potential studies. 1560 were excluded after abstract screening. 136 reports were assessed for eligibility, with a further 112 excluded (Figure 1). The updated literature search on 31^st^ January 2023, returned a further 139 abstracts. Of these, 133 were excluded upon abstract screening and 6 reports were assessed for eligibility, with a further 5 excluded. The most common assessment methods employed by included studies (17/25, 68%) were postmortem neuropathological changes, followed by hippocampal volume studies (5/25, 20%), and CSF studies of AD-biomarkers (3/25; 12%) (Table 1). Postmortem studies mainly focussed on the presence of AD-related pathology, i.e., either amyloid plaques, NFTs, or both, although several postmortem studies also investigated a range of other markers, including histopathological changes (e.g., features of Parkinson’s and Lewy body disease, vascular pathology), protein expression, and the presence of various peptides and ions. Where authors had included statistical comparisons between groups, we included these in Table 1. For relevant descriptive findings, we have included these in the notes to Table 1.

**Figure 1:**
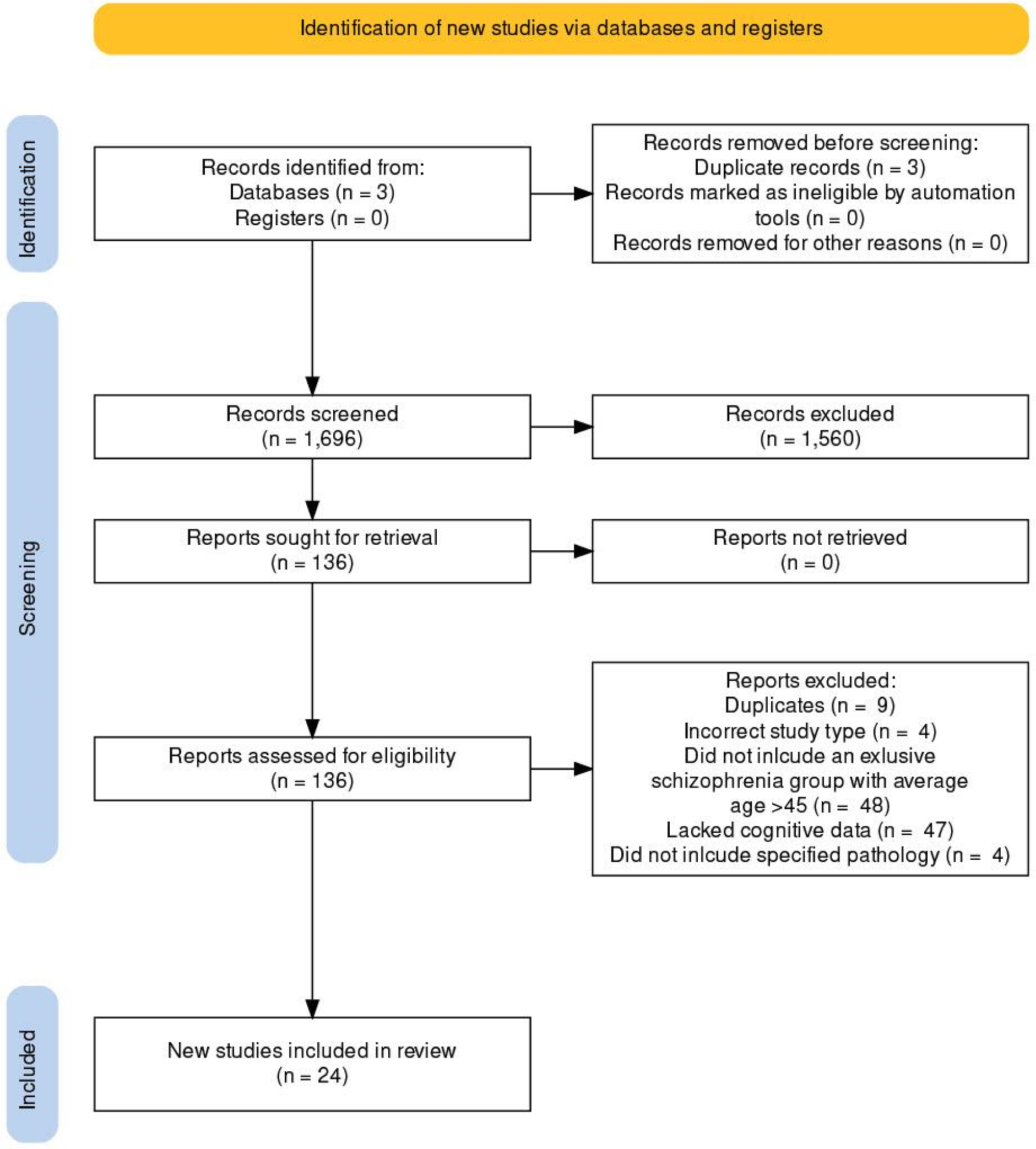
PRISMA flowchart.

**Figure 2:**
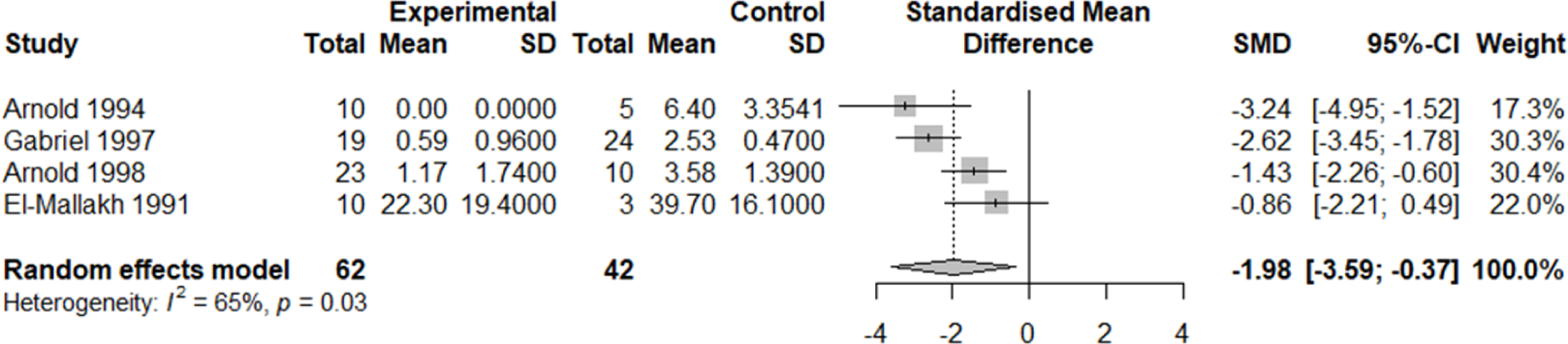
Number of Amyloid Plaques in Schizophrenia vs Alzheimer’s. Forest plot showing the effect size (Hedge’s G) and the pooled effect size for studies comparing APs in the schizophrenia group vs AD. ‘Total’ denotes the sample size, SD denotes standard deviation, SMD denotes standardised mean difference 95% CI denotes 95% confidence interval.

**Figure 3.**
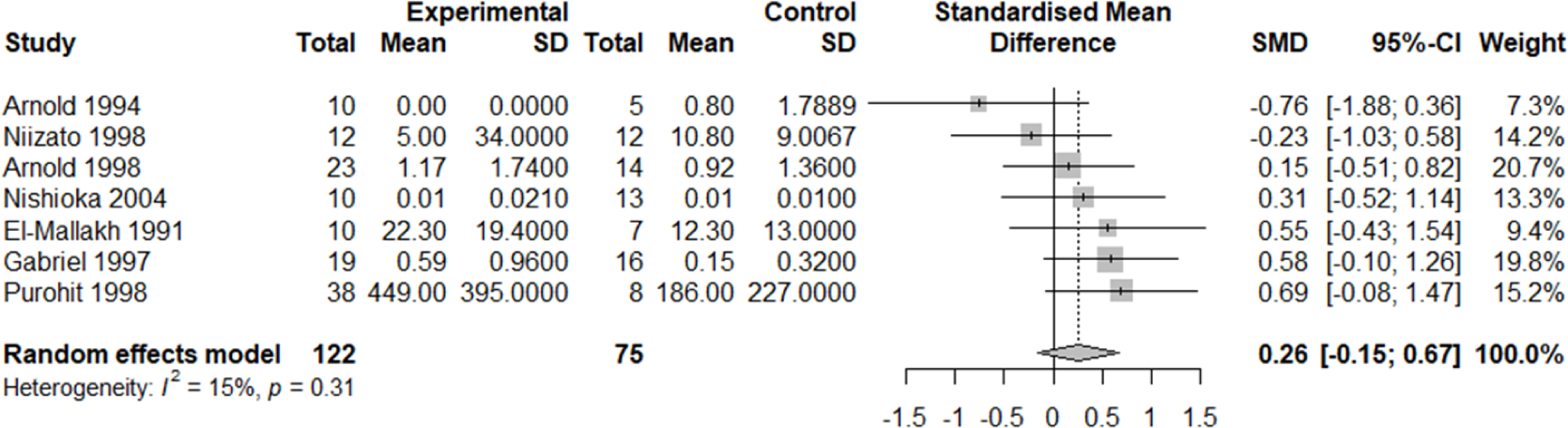
Number of Amyloid Plaques in Schizophrenia vs Controls. Forest plot showing the effect size (Hedge’s G) and the pooled effect size for studies comparing APs in the schizophrenia group vs controls. ‘Total’ denotes the sample size, SD denotes standard deviation, SMD denotes standardised mean difference 95% CI denotes 95% confidence interval.

**Figure 4.**
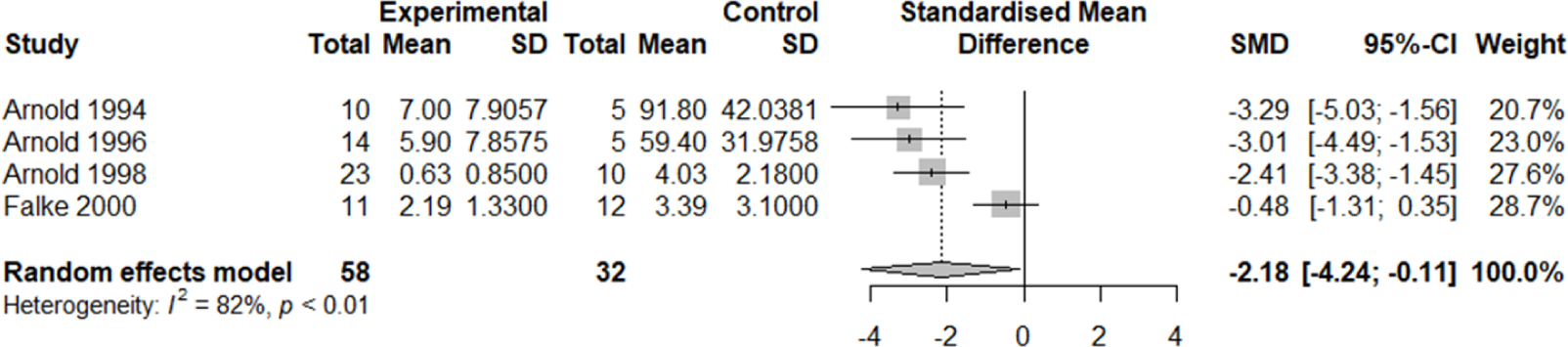
Number of Neurofibrillary Tangles in Schizophrenia vs Alzheimer’s. Forest plot showing the effect size (Hedge’s G) and the pooled effect size for studies comparing NFTs in the schizophrenia group vs AD. ‘Total’ denotes the sample size, SD denotes standard deviation, SMD denotes standardised mean difference 95% CI denotes 95% confidence interval.

**Figure 5.**
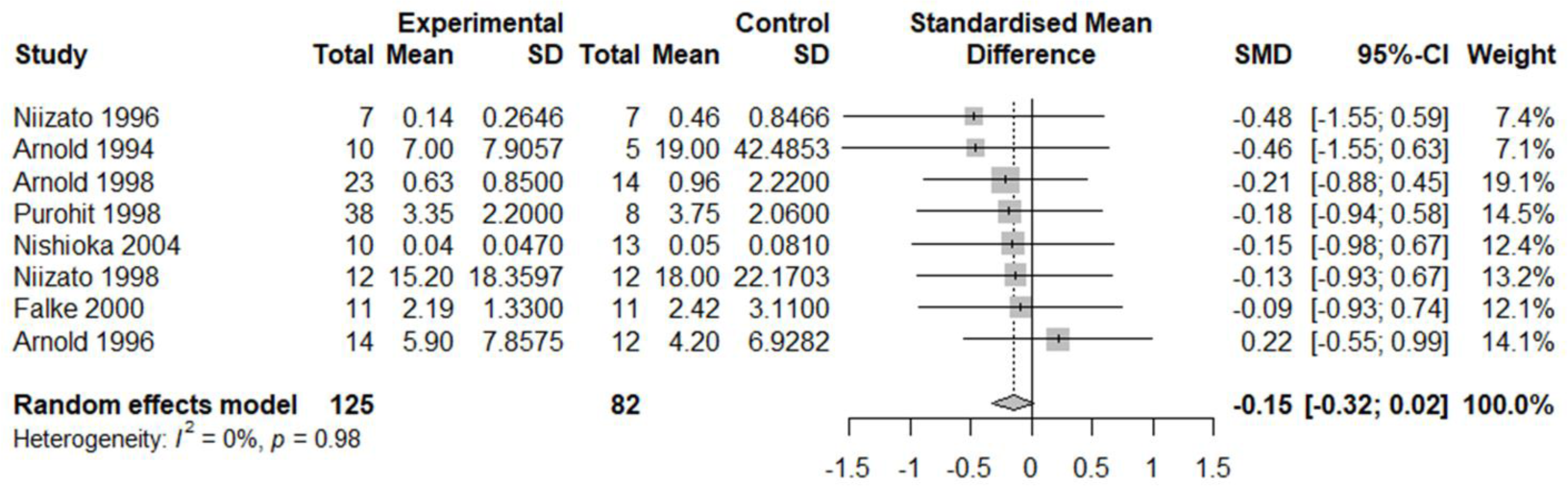
Number of Neurofibrillary Tangles in Schizophrenia vs controls. Forest plot showing the effect size (Hedge’s G) and the pooled effect size for studies comparing NFTs in the schizophrenia group vs controls. ‘Total’ denotes the sample size, SD denotes standard deviation, SMD denotes standardised mean difference 95% CI denotes 95% confidence interval.

### Neurodegeneration marker findings

In terms of AD-related pathology on postmortem examination, no studies found a significant increase of amyloid plaques or NFTs in cognitively impaired schizophrenia patients compared to controls. Nine postmortem studies compared amyloid-β plaques (APs) or NFTs in people with schizophrenia to an additional AD control group; all of these studies found significantly more APs or NFTs in the AD group compared to the schizophrenia group (Table 1). Some authors included other AD-related markers, including acetylcholinesterase activity,[20] Alz-50 immunoreactivity (a marker for Tau),[21] and the number of neurons that stained positive for the presence of tau[22]. In each of these studies, the cognitively impaired schizophrenia group did not differ significantly from the normal control group, and showed significantly lower levels of the AD pathology compared to an AD control group.

Decreases in CSF concentrations of amyloid-beta 42 (Aβ42) and elevations in total tau and phosphorylated tau are used as biomarkers for AD.[23] Two studies that measured CSF Aβ42 levels found significantly lower levels of Aβ42 in schizophrenia patients compared to normal controls (Table 1). However, these studies also found Aβ42 levels in schizophrenia patients to be significantly lower compared to the AD comparison group.[24, 25] AD patients also had significantly elevated levels of T-tau and P-tau compared to schizophrenia patients. No significant differences in CSF T-tau or P-tau levels were found between schizophrenia patients and controls in these studies, or in an additional study that only looked at T-tau and P-tau in schizophrenia patients vs controls.[26].

Studies that investigated hippocampal MRI volume differences have reported inconsistent findings. One study comparing treatment resistant patients with schizophrenia (TRS) and non-treatment resistant patients with schizophrenia (NTRS) found significantly smaller hippocampal volumes in TRS compared to controls but no difference between NTRS and controls.[27] A further study did not find any significant difference in hippocampal volume between schizophrenia patients and controls, but did note that there was a steeper decline in hippocampal volume as a function of age in the schizophrenia group. [28] One study found smaller left sided hippocampal-amygdala volumes for schizophrenia versus controls.[29] Rivas et al compared schizophrenia patients who showed signs of dementia with schizophrenia patients who did not, and controls. The schizophrenia group with signs of dementia had significantly lower mean whole hippocampal volumes compared to the healthy control group, while the schizophrenia patients without cognitive impairment showed no significant difference compared to healthy controls. Differences for whole hippocampal volume between the schizophrenia group with dementia and the schizophrenia group without dementia were not significant.[30] Prestia et al found cognitively impaired schizophrenia patients had similar hippocampal volumes to AD patients, and significantly lower volumes than controls.[31]

### Meta analysis findings

In total, 10 studies were deemed suitable for metaanalysis. Details of why studies were excluded from the meta-analysis are presented in supplementary table 1. The pooled effect size for included studies comparing APs in the schizophrenia group vs AD was −1.98 (−3.59; −0.37) and the pooled effect size for included studies comparing NFTs in the schizophrenia group vs AD was −2.18 (−4.24; −0.11). The pooled effect size for included studies comparing APs in the schizophrenia group vs controls was 0.26 (−0.15; 0.67) and the pooled effect size for included studies comparing NFTs in the schizophrenia group vs controls was −0.15 (−0.32; 0.02). Thus, the pooled effect size was significantly lower in schizophrenia patients vs AD for both APs and NFTs. There was no significant difference in pooled effect size between schizophrenia patients and controls for either APs or NFTs.

## Discussion

Impaired cognition is a core symptom of schizophrenia that precedes the onset of psychosis, is a key determinant of poor functioning, and is the most resistant to treatment. It remains unclear whether cognition worsens subsequent to diagnosis.[32,33] Recent studies have suggested that there is an additional modest decline in cognitive functioning in later life in people with schizophrenia (though this decline is heterogenous, and detection is dependent on the cognitive measures used),[7,34] and several population-based studies have identified significantly higher rates of dementia diagnoses in schizophrenia.[1,3–5,35,36] These findings persist when controlling for dementia risk factors (e.g. smoking, diabetes) and taking into account incentives to diagnose dementia (e.g. obtaining admission to nursing homes). We lack a good understanding of the underlying cause of cognitive decline in schizophrenia.

Our systematic review shows that published studies have not found higher rates of dementia pathologies, specifically AD-related pathology, in cognitively impaired schizophrenia individuals compared to controls. How does this conclusion sit with the consistent finding of increased rates of dementia generally and AD specifically diagnosed in population studies?[1,35–37] One potential explanation is that there is a neurodegenerative process intrinsic to schizophrenia, which is not detectable with standard neuropathological marker examination.[38] For example, some abnormal neurodevelopmental processes (e.g. relating to synaptic plasticity) may continue throughout life.[39] Individuals with schizophrenia also have increased risk factors (e.g., higher rates of drug and alcohol use, socioeconomic adversity, and antipsychotic use) that impact cognition, which may not cause macroscopic pathological changes identifiable at post-mortem examination. Another possible explanation is that normal ageing processes have a more notable effect on patients with schizophrenia due to diminished cognitive reserve, which has implications for the clinical assessment and diagnosis of dementia in these individuals.

The cognitive assessment tools used to clinically diagnose dementia are potentially insufficiently sensitive to differentiate between dementia diseases and schizophrenia-related cognitive impairment. Positive and negative symptoms of schizophrenia could reduce scores on commonly used tests through distraction, reduced concentration, memory and fluency, so that patients with schizophrenia will score lower than the general population on cognitive testing, regardless of the age when they are tested. Given their lower starting point, even mild drops in test scores (for instance associated with normal ageing), could place a patient below the cut-off used to diagnose dementia.

In addition to cognitive testing, assessments for dementia usually include brain imaging. Clinicians use MRI and CT to look for signs of vascular disease and hippocampal atrophy, which is suggestive of AD. The studies we identified that quantified hippocampal volume showed mixed results. One showed this to be significantly lower than controls and similar to AD patients. It does seem that some patients with schizophrenia have reduced hippocampal volumes. However, the average age in these studies was lower than would be expected for AD and reduced hippocampal volume is consistently reported in schizophrenia patients in the absence of diagnosed dementia, and early in the disease.[40–42] It seems more likely that reduced hippocampal volume is a primary feature of the disease, rather than secondary to comorbid dementia. Reduced hippocampal volume may however increase diagnostic uncertainty when assessing a patient with schizophrenia for dementia, as it could mimic the appearance of hippocampal atrophy seen in AD.

Our analysis does not shed light on which of these non-mutually exclusive factors may be the primary driver of declining cognition in later life in schizophrenia. Further research on AD-related biomarkers such as PET and/or CSF amyloid and tau measures, markers of vascular dementia, PD, LBD and other measures of neurodegeneration (e.g. neurofilament light chain) may help to confirm the true rate of AD and other dementias in the schizophrenia population. Future studies are also needed to further characterise the trajectory of cognitive decline in schizophrenia patients who have been diagnosed with comorbid dementia, in comparison to the trajectory of normal ageing and dementia. A trajectory which appears distinct from either of these may be suggestive of a neurodegenerative process intrinsic to schizophrenia. Indeed, greater clinical use of cognitive assessments in the schizophrenia population would not only provide richer pools of data for research, but may also better characterise patient functioning and needs.[7].

### Limitations

The higher rates of diagnosed dementia have been proposed to be related to the effects of vascular risk factors or antipsychotic medication.[43] Only two studies in our review[44, 45] specifically looked for vascular pathology, and no studies looked specifically at medication naive patients, which limits our ability to make conclusions about these potential dementia risk factors. In the wider literature the picture for antipsychotic use is mixed, with some studies showing they *reduce* the risk of dementia,[5] and that untreated psychosis is linked to worsening cognition.[46] With regards to vascular pathology, the two studies included in our review did not find significantly higher rates than what would be expected in the general population, and population studies have also attempted to control for vascular risk factors.

A number of studies that investigated post-mortem changes in schizophrenia were not included in our study, as they did not specifically investigate patients who had established dementia or cognitive decline. Out of these, there were a handful of early studies that did in fact identify increased rates of AD in patients with schizophrenia based on neuropathological criteria.[47,48] However, these studies were performed prior to the widespread use of formalised criteria required today. Following the advent of such criteria, these results were not replicated. In fact, older schizophrenia populations were found to have equal to or less pathology compared to elderly controls.

Our analysis was restricted to studies which examined patients who were reported to have dementia or had evidence of cognitive impairment. Our analysis was also limited to studies that included a comparison group. However, other post-mortem studies that have looked at all patients with schizophrenia regardless of cognition have reported the same finding; no higher rates of AD pathology compared to that in the general population.[49–51].

Finally, the majority (14/17) of included post-mortem studies were completed between 1990 and 2000, and before the publication of the National Institute for Ageing and the Alzheimer’s Association (NIA-AA) research framework which defined in vivo biological criteria for AD, which included measures of aggregated Aβ and tau in the CSF.[52] Thus the AD pathological assessments reported in the included studies do not fully align with current AD diagnostic criteria. The three studies in our review which analysed CSF for AD biomarkers did not report findings consistent with AD pathology, and none found any significant differences in Total Tau and pTau between controls and schizophrenia patients. Two studies did show lower levels of Aγ1–42 in schizophrenia patients compared to controls, but these levels were still significantly higher than the levels seen in the AD comparison groups.

## Conclusion

Overall, we have shown that the observed increased rate of dementia in patients with schizophrenia is unlikely to be attributable to a higher rate of common neurodegenerative processes such as AD. Our findings highlight potential limitations in the etiological diagnosis of dementia in older individuals with schizophrenia, and the need for further investigation on what underlies the relatively modest reported declines in cognition with age.

## Data Availability

All data produced in the present study are available upon reasonable request to the authors

## Acknowledgements

RH is supported by University College London Hospitals’ National Institute for Health Research (NIHR) Biomedical Research Centre. KL is funded by the UK Medical Research Council (MR/S021418/1).

## Contributorship Statement

Jack C. Wilson: Planning, abstract screening, data extraction, writing report. Kathy Y. Liu: Planning, abstract screening, editing report Katherine Jones: Abstract screening, data extraction Jansher Mahmood: Abstract screening, data extraction Utkarsh Arya: Planning, abstract screening, data extraction Robert Howard: Planning, editing report

↔ No significant difference

↑ Significantly higher

↓ Significantly lower<colcnt=1>

**+** Lewy bodies, ubiquinated dystrophic neurites, GFAP in astrocytes, resting and active microglia

**+ +** Cerebral Amyloid Angiopathy, Lewy bodies, vascular pathology such as infarcts, atherosclerosis, arteriolar sclerosis, Features of Parkinson’s disease, diffuse Lewybody disease, Creutzfeldt-Jakob disease, and Pick’s disease.

**+ + +** Cerebral Amyloid Angiopathy, Lewy bodies, vascular pathology such as infarcts, atherosclerosis, arteriolar sclerosis, neocortical neuronal loss, hirano bodies, neuropil degeneration and gliosis; hippocampal degeneration, granulovacuolar degeneration

* Study also included 7 Schizophrenia patients without cognitive impairment, which we have not included

** No evidence of morphological features of other dementing neurodegenerative conditions, such as multi-infarct dementia, Parkinson’s disease, diffuse Lewy-body disease, Creutzfeldt-Jakob disease, or Pick’s disease, was found

***Postmortem neuropathologic examination findings: Alzheimer’s disease: 9, Parkinson’s disease: 2, Multi infarct dementia: 1, Multiple sclerosis: 1, Ischaemic cerebrovascular disease 23, Secondary neoplasms 3, Brainstem Haemorraghe: 1, parietal lobe haemangioma: 1, Frontal Leukotomy: 11

**** NFTs were only present in samples from the AD group

*****Steeper decline as function of age in Sz group

****** Several other Aβ peptides also lower

## Key

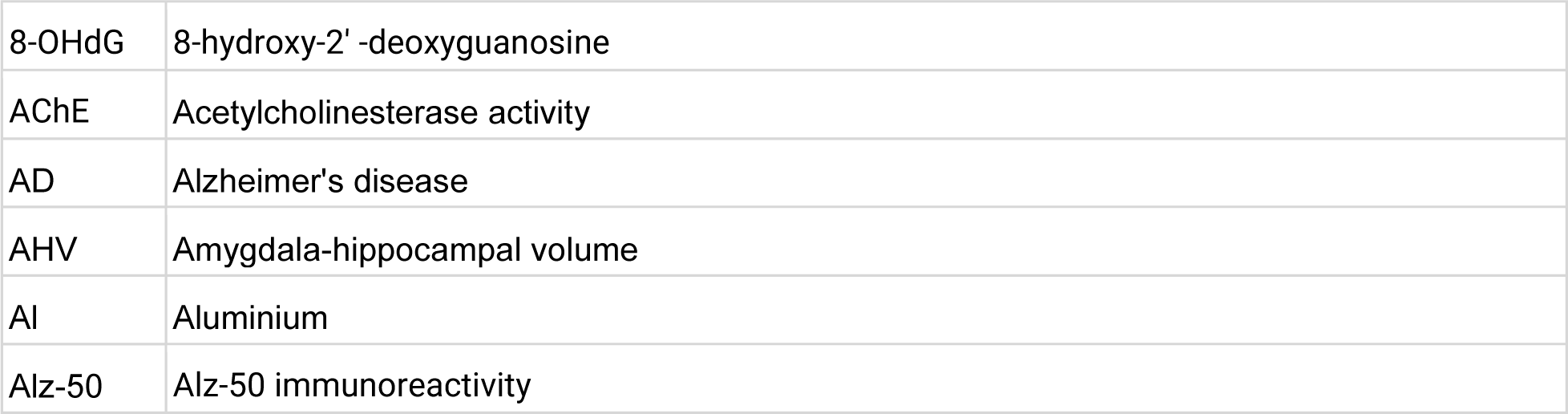
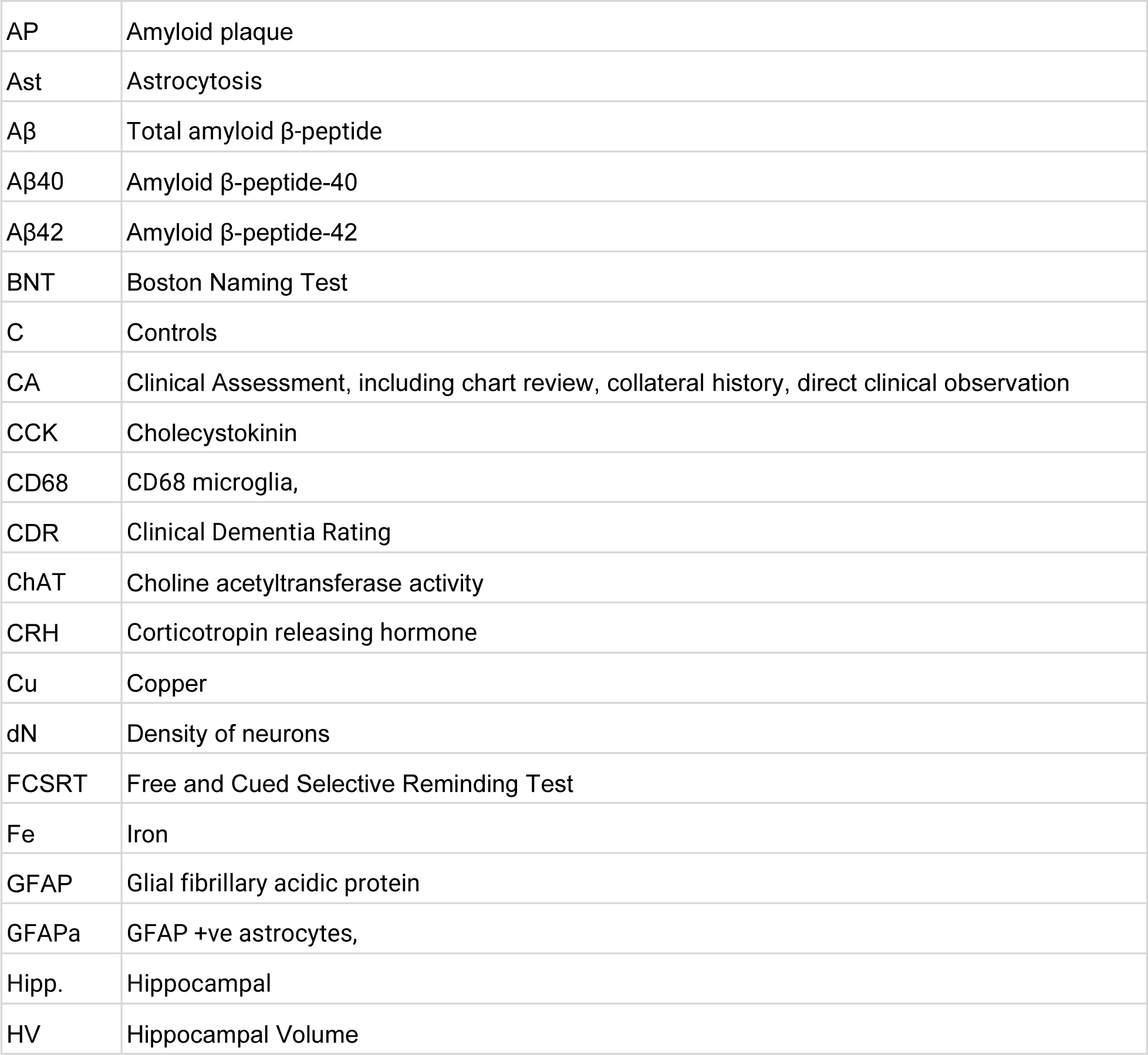
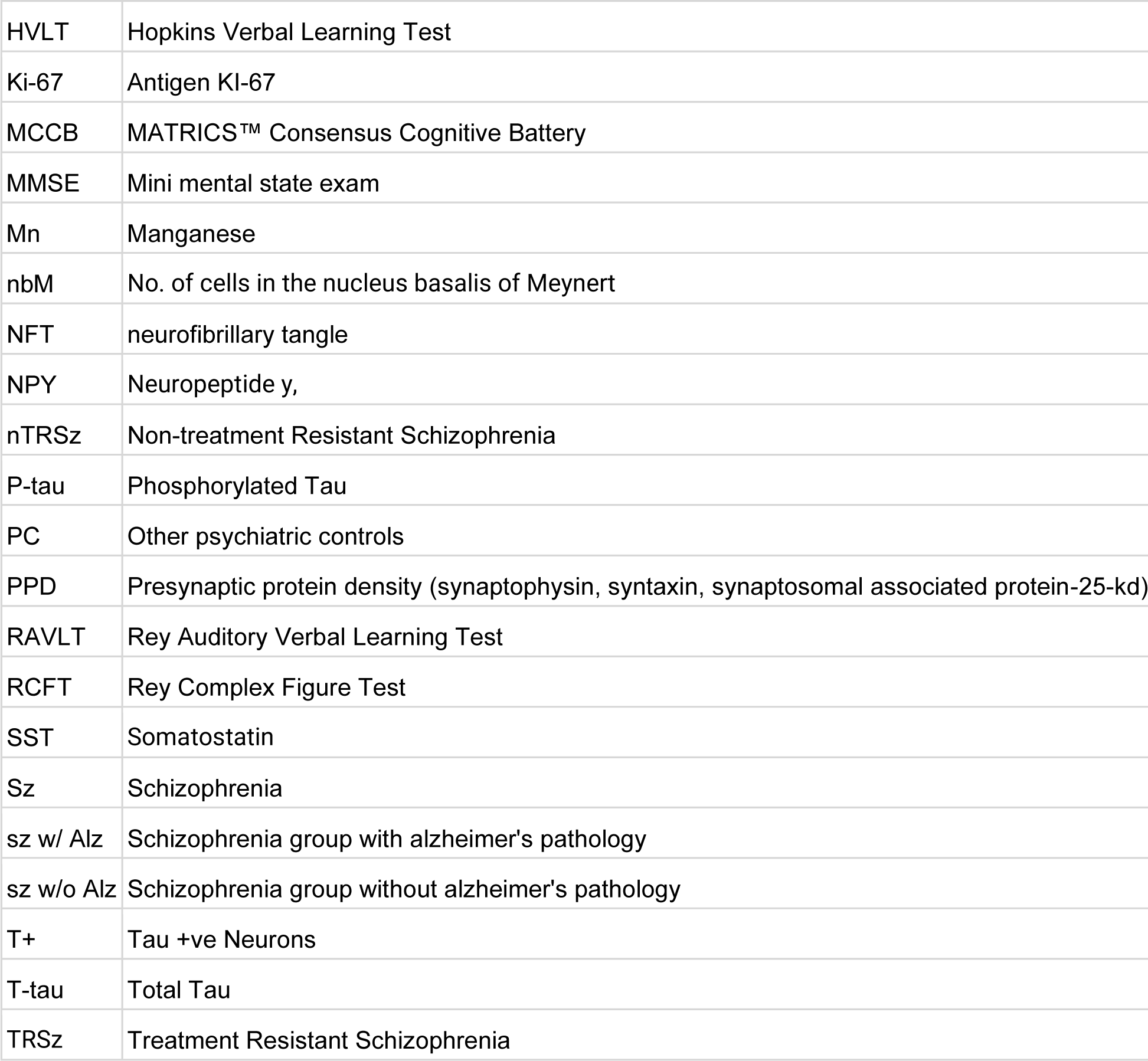
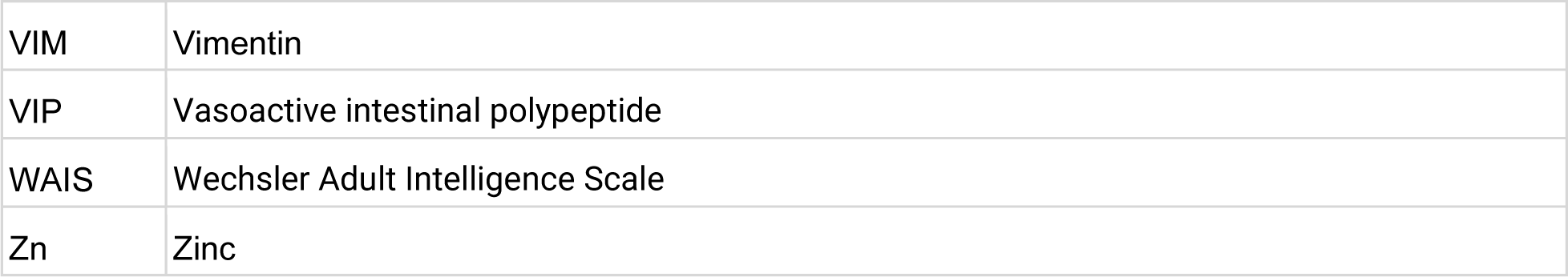

---

**Supplementary Table 1:**
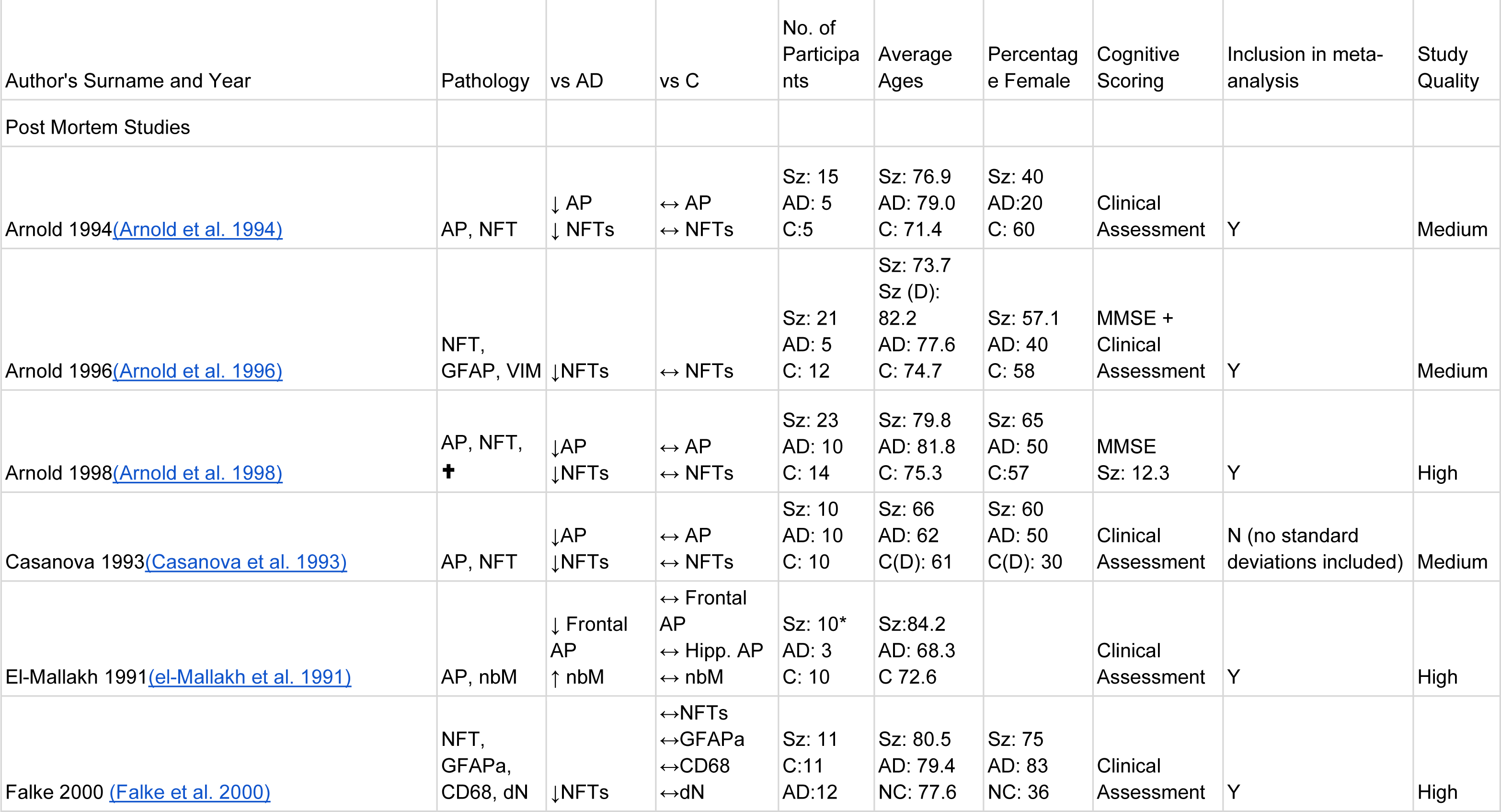
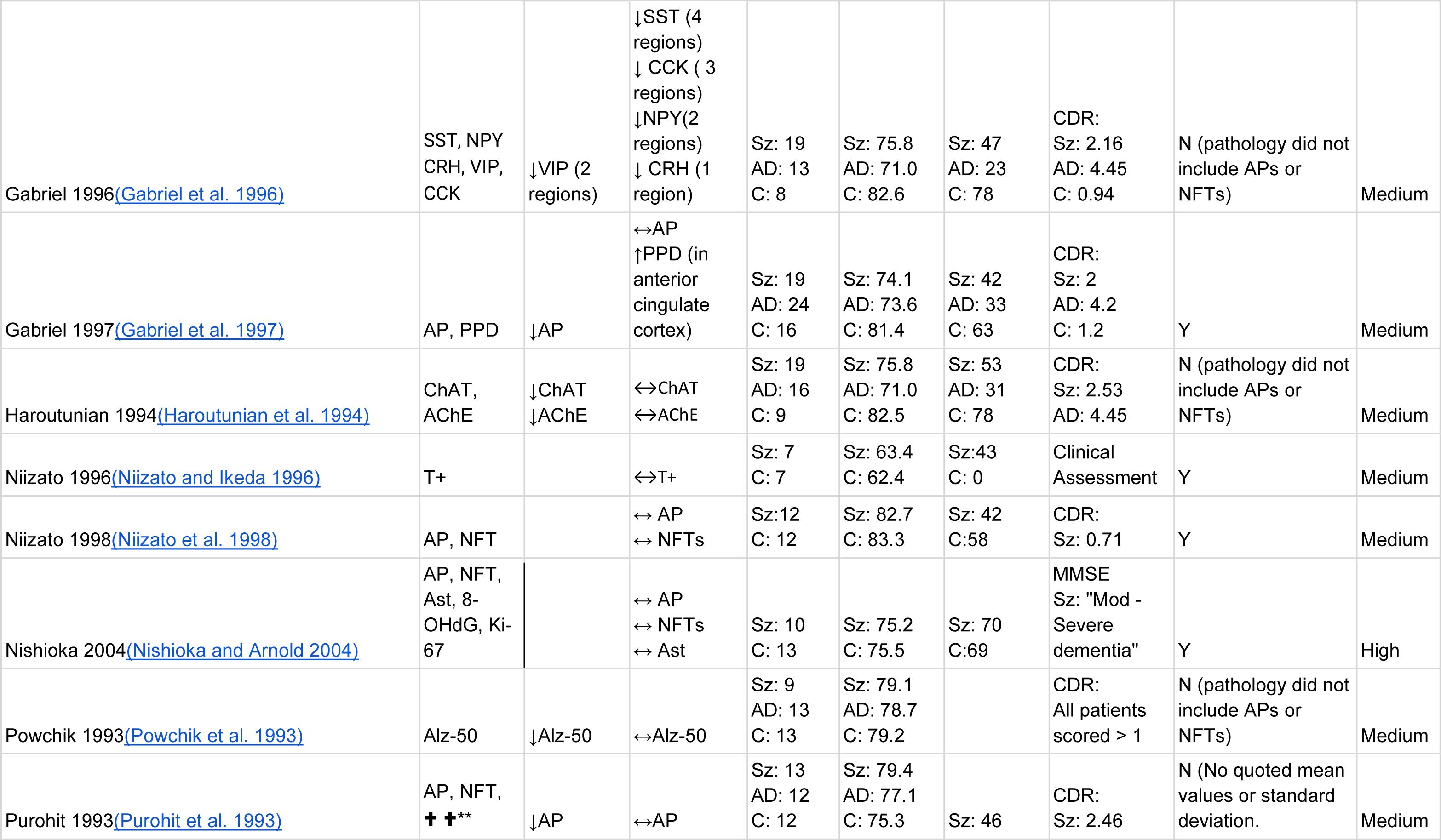
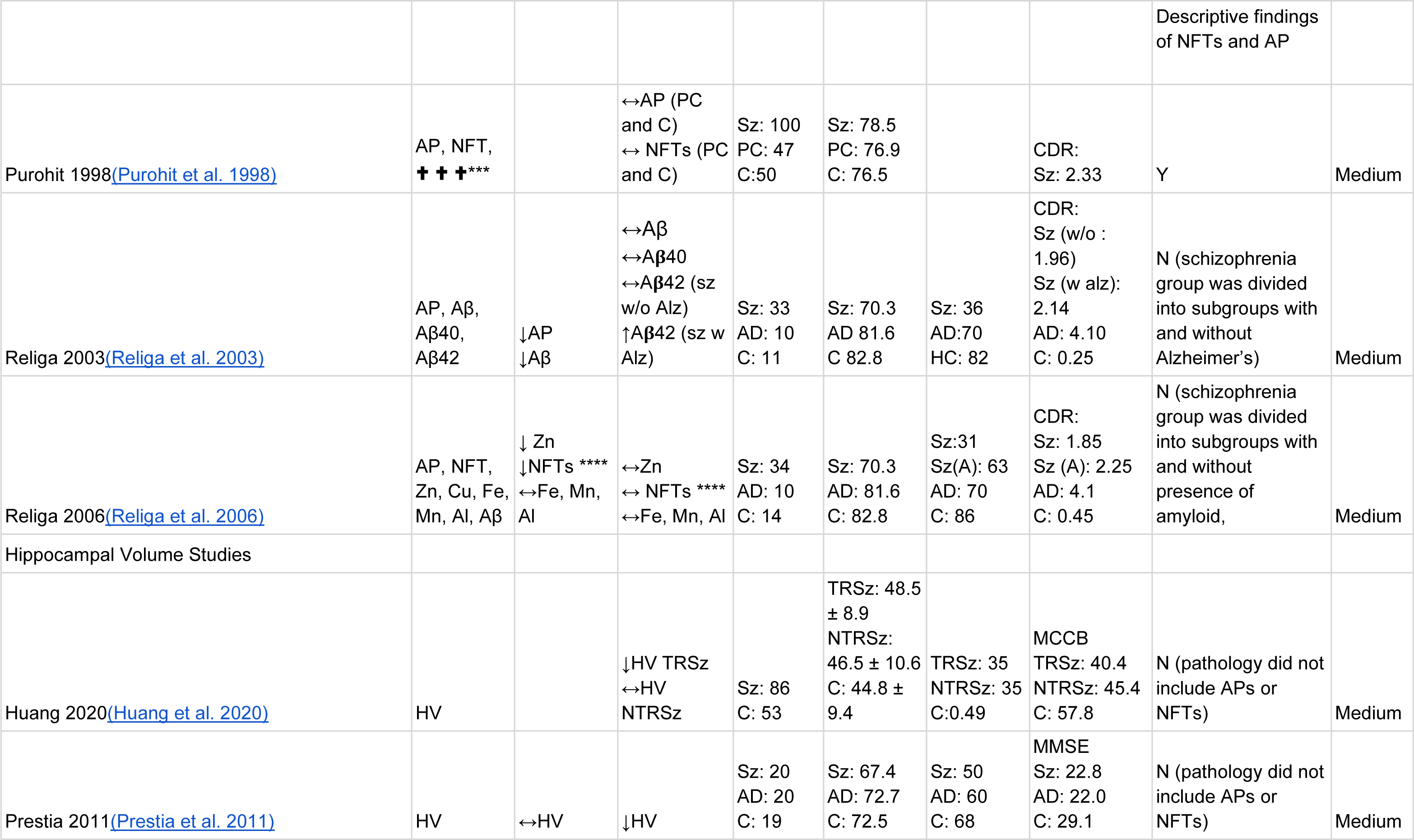
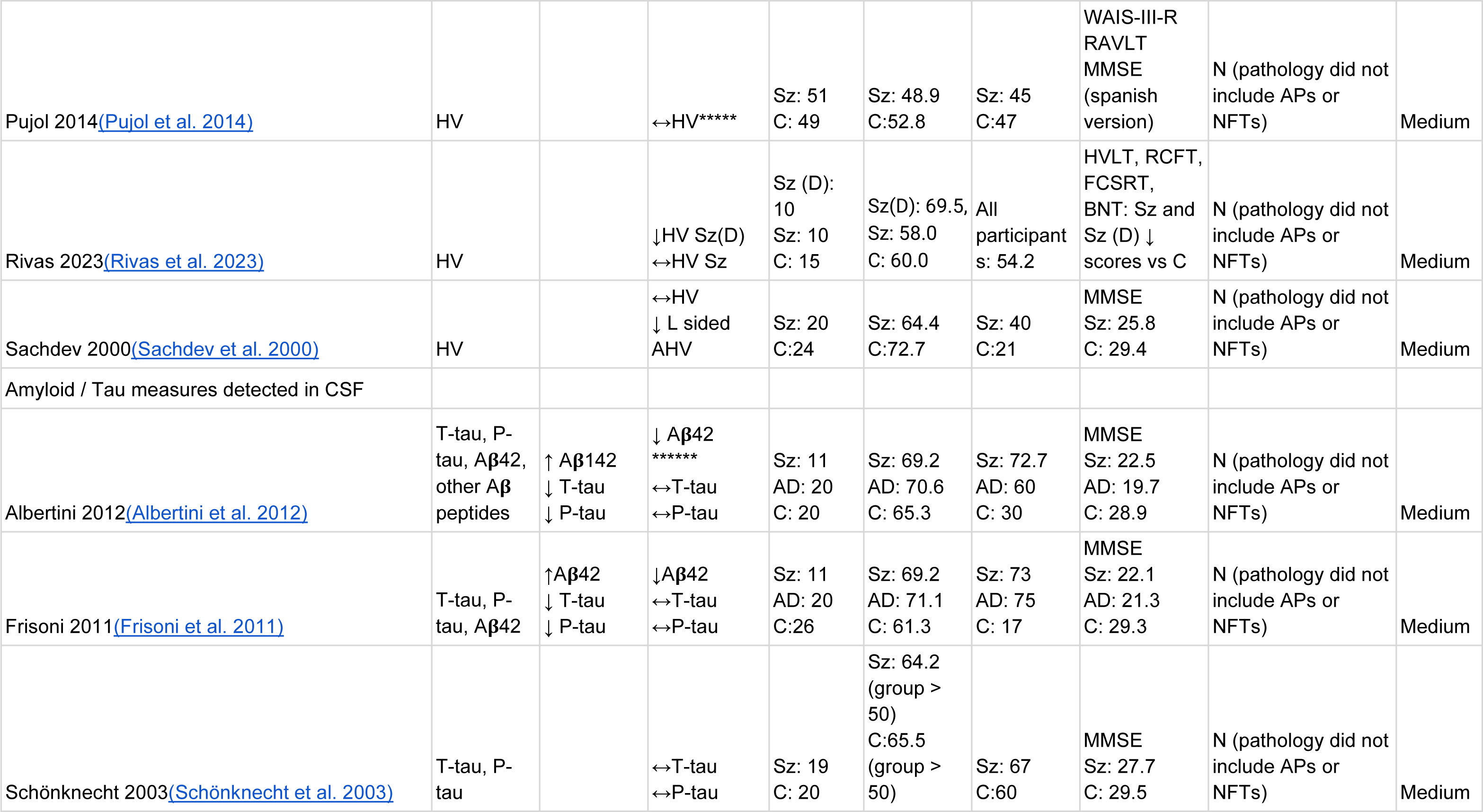
Characteristics and Findings of included studies. Quality assessment was performed using a modified version of the NIH Quality Assessment Tool for Observational Cohort and cross sectional studies. Studies were given a score of between 0 and 12, with 0-4 classed as low, 5-8 classed as medium, and 9-12 classed as High.

